# Who should we test for COVID-19? A triage model built from national symptom surveys

**DOI:** 10.1101/2020.05.18.20105569

**Authors:** Saar Shoer, Tal Karady, Ayya Keshet, Smadar Shilo, Hagai Rossman, Amir Gavrieli, Tomer Meir, Amit Lavon, Dmitry Kolobkov, Iris Kalka, Anastasia Godneva, Ori Cohen, Adam Kariv, Ori Hoch, Mushon Zer-Aviv, Noam Castel, Caroel Sudre, Anat Ekka Zohar, Angela Irony, Tim Spector, Benjamin Geiger, Dorit Hizi, Varda Shalev, Ran Balicer, Eran Segal

## Abstract

The gold standard for COVID-19 diagnosis is detection of viral RNA in a reverse transcription PCR test. Due to global limitations in testing capacity, effective prioritization of individuals for testing is essential. Here, we devised a model that estimates the probability of an individual to test positive for COVID-19 based on answers to 9 simple questions regarding age, gender, presence of prior medical conditions, general feeling, and the symptoms fever, cough, shortness of breath, sore throat and loss of taste or smell, all of which have been associated with COVID-19 infection. Our model was devised from a subsample of a national symptom survey that was answered over 2 million times in Israel over the past 2 months and a targeted survey distributed to all residents of several cities in Israel. Overall, 43,752 adults were included, from which 498 self-reported as being COVID-19 positive. We successfully validated the model on held-out individuals from Israel where it achieved a positive predictive value (PPV) of 46.3% at a 10% sensitivity and demonstrated its applicability outside of Israel by further validating it on an independently collected symptom survey dataset from the U.K., U.S. and Sweden, where it achieved a PPV of 34.7% at 10% sensitivity. Moreover, evaluating the model’s performance on this latter independent dataset on entries collected one week prior to the PCR test and up to the day of the test we found the highest performance on the day of the test. As our tool can be used online and without the need of exposure to suspected patients, it may have worldwide utility in combating COVID-19 by better directing the limited testing resources through prioritization of individuals for testing, thereby increasing the rate at which positive individuals can be identified and isolated.

## Introduction

The rapid and global spread of COVID-19 led the World Health Organization (WHO) to declare it a pandemic on March 11, 2020. One major factor that contributes to the spread of the virus is the apparently large number of undiagnosed infected individuals. This knowledge gap facilitates the silent propagation of the virus, delays the response of public health officials and results in an explosion in the number of cases (1,2).

One reason for this knowledge gap is insufficient testing. While the current gold standard for COVID-19 diagnosis is detection of viral RNA in a reverse transcription PCR test, the number of tests is limited by financial and logistic constraints. In a time when almost all countries are faced with the same health challenge, resources are scarce. This creates the need for a prioritization mechanism to allocate tests and resources more efficiently towards individuals who are more likely to test positive, leading to earlier identification of COVID-19 patients and reduced spread of the virus. Despite this need, most countries still employ a simplistic testing strategy based on display of symptoms associated with the disease and either close epidemiological contact with a confirmed COVID-19 case or belonging to a high risk group(3). In practice, these strategies lead to a relatively small fraction of positive tests among those tested and thus to inefficient use of the precious testing resources.

Here, we present a model that provides statistically significant estimates of the probability of an individual to test positive for SARS-CoV-2 infection in a PCR test, based on a national symptom survey that we distributed in Israel. Notably, while most studies describing the clinical characteristics of COVID-19 cases were based on symptoms of hospitalized patients (4), our survey data allowed us to also study symptoms of milder cases and reveal which symptoms hold the highest predictive power for COVID-19 diagnosis. Using our model, the risk for a positive COVID-19 test can be evaluated in less than a minute and without added costs, or risk of exposure to a suspected patient. Our approach can thus be used globally to make more efficient use of available testing capacities, by significantly increasing the fraction of positive tests obtained, and by rapidly identifying individuals that should be isolated until definitive test results are obtained.

## Methods

### Data

In this study, we utilized data that originates from two versions of a one-minute survey that was developed and deployed by our research group in the early stages of the COVID-19 spread in Israel (5). The online version of the survey includes questions relating to age, gender, prior medical conditions, smoking habits, self-reported symptoms and geographical location (see section 1a in the supplementary appendix). Questions regarding prior medical conditions and symptoms included in the survey were carefully chosen by medical professionals. Each participant is asked to fill the survey once a day for himself and for family members that are unable to fill it for themselves (e.g. children and the elderly). The survey is anonymous to maintain the privacy of the participants, and has been collected since March 14th, 2020. As the number of COVID-19 diagnosed individuals in Israel rose, in some cities more than others, a shortened version of the survey was deployed using an Interactive Voice Response (IVR) platform. This version of the survey included information on respondents’ age group, gender, presence of prior medical conditions, general feeling and a partial list of symptoms, including fever, cough, shortness of breath, sore throat and loss of taste or smell (see Section 1b in the supplementary appendix). Cities were targeted to participate in the IVR version of the survey according to the number of diagnosed patients and an increased concern for COVID-19 outbreaks (Table S1). Starting April 5, 2020, citizens in the targeted cities were contacted and responses were collected anonymously. This study followed the Transparent Reporting of a Multivariable Prediction Model for Individual Prognosis or Diagnosis (TRIPOD) reporting guideline (6).

### Study Design and Population

Overall, 695,586 and 66,447 responses were collected up until April 26th, 2020, from the online and IVR versions of the survey, respectively. Since children express different clinical manifestations of COVID-19 infection (7,8), we decided to focus our analysis only on adults (age above 20 years old). To avoid translation discrepancies, we included only responses in Hebrew. We also excluded responses that did not meet quality control criteria, such as a reasonable age (<120 years old) and body temperature (35-43°C) or responses suspected as spam (see Section 3 in the supplementary appendix) (Figure 1). A total of 33,737 responses were eventually included from the IVR version of the survey. Since surveyed cities were at relatively high risk (Table S1), the prevalence of COVID-19 diagnosed responders in the IVR data was 1.14%, 6 times higher than the national prevalence of 0.18% (3). These cities also had very high response rates, between 6% to 16% of the cities’ population (Table S1). From the online version of the survey, we randomly sampled a single response for each individual that was recognized by a unique identifier we created (see Section 4 in the supplementary appendix), and when an individual reported a COVID-19 diagnosis, we randomly sampled one response only from those which included a positive diagnosis answer. A total of 131,166 responses were identified in the online version, of which 0.09% reported a positive COVID-19 diagnosis, which is closer to the national prevalence of 0.18% (3). The characteristics of these unique responders are described in Table S2 of the supplementary appendix.

**Figure 1.**
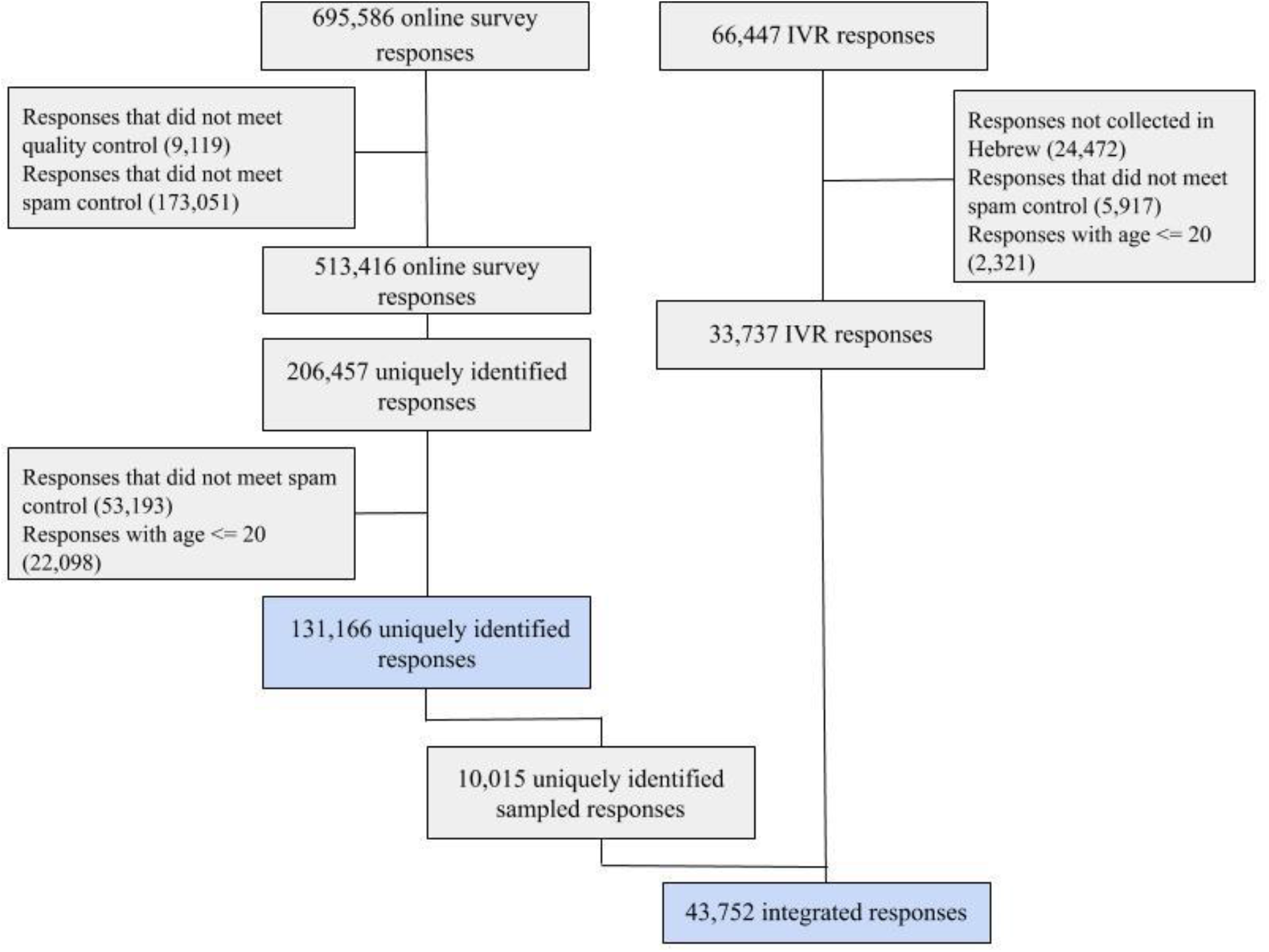
Study population flow chart. Numbers represent recorded responses. Blue colored boxes show reponses which were used in extended features model (top) and primary model (bottom) constructions.

For the integration of the two survey versions, all 33,737 IVR responses were combined together with all 114 uniquely identified responders in the online survey that self-reported COVID-19 diagnosis and a random sample of 9,901 undiagnosed responders in the online version, to maintain the same diagnosis prevalence as in the IVR version (Figure 1). Overall, 43,752 responses were eventually included in the study, of which 498 self-reported as being COVID-19 diagnosed. The characteristics of these responders are described in Table 1.

**Table 1.** Baseline characteristics of the primary model population.

| **Table 1. Baseline characteristics of the primary model population** |  |  |  |  |  |
| --- | --- | --- | --- | --- | --- |
| **Characteristic,**<br>**mean (SD) or %** | **All individuals**<br>**n=43,752**<br>**(100%)** | **IVR version**<br>**n=33,737**<br>**(77.11%)** | **Online version**<br>**n=10,015**<br>**(22.89%)** | **COVID-19**<br>**undiagnosed**<br>**n=43,254**<br>**(98.862%)** | **COVID-19**<br>**diagnosed**<br>**n=498**<br>**(1.138%)** |
| Age in years | 44.941<br>(15.499) | 43.897<br>(15.244) | 48.460<br>(15.831) | 44.894<br>(15.47) | 49.076<br>(17.363) |
| Gender - male | 23,630<br>(54.009%) | 19,151<br>(56.766%) | 4,479<br>(44.723%) | 23,339<br>(53.958%) | 291<br>(58.434%) |
| COVID-19 diagnosed | 498 (1.138%) | 384 (1.138%) | 114 (1.138%) | 0 (0.0%) | 498 (100.0%) |
| Prior medical<br>conditions | 8,070<br>(18.943%) | 5,176<br>(15.884%) | 2,894<br>(28.897%) | 7,946<br>(18.861%) | 124<br>(26.271%) |
| **Symptoms** |  |  |  |  |  |
| Feel well | 41,661<br>(95.221%) | 32,132<br>(95.243%) | 9,529<br>(95.147%) | 41,217<br>(95.291%) | 444<br>(89.157%) |
| Sore throat | 1,507 (3.445%) | 1,141 (3.382%) | 366 (3.655%) | 1,422 (3.288%) | 85 (17.068%) |
| Cough | 2,138 (4.887%) | 1,459 (4.325%) | 679 (6.78%) | 1,984 (4.587%) | 154 (30.924%) |
| Shortness of breath | 576 (1.317%) | 443 (1.313%) | 133 (1.328%) | 505 (1.168%) | 71 (14.257%) |
| Loss of taste or smell | 605 (1.388%) | 545 (1.624%) | 60 (0.599%) | 469 (1.088%) | 136 (27.812%) |
| Fever (body<br>temperature above 38<br>°C) | 77 (0.176%) | 53 (0.157%) | 24 (0.24%) | 64 (0.148%) | 13 (2.61%) |

### Predicting the Outcome of a COVID-19 Test

We defined survey self-reporting of a COVID-19 laboratory confirmed diagnosis as our outcome. We constructed two models. The first, which we term the primary model, was constructed from the integrated responses from both the IVR and online versions of the survey and included the reduced set of questions that were surveyed in the IVR version. The second model, which we term the extended features model, was constructed using only responses from the online version, and included additional symptoms and questions that were not part of the IVR survey. We trained both the primary and extended features models using Logistic Regression. In addition, in order to capture nonlinear interactions and interactions amongst features, in both cases we also constructed models using a Gradient Boosting Decision Trees algorithm (9). To test the validity of our models, all model constructions were done using the framework of cross-validation, in which model performance is evaluated on a subset of the data that was not used in the model’s construction. For more information on handling of missing values and the models construction process, see Section 6 in the supplementary appendix.

### Primary Model

The primary model was constructed using responses to both the IVR and the online version of the survey. Features included in this model were determined by the IVR version, since it included a subset of the online version questions. These consisted of age group, gender, presence of prior medical conditions, general feeling, and the following symptoms: fever, cough, shortness of breath, sore throat and loss of taste or smell.

### Extended Features Model

The extended features model was constructed using only responses from the online version of the survey, as it had 14 additional features that were not available in the IVR version. This extended list added dry cough and moist cough (instead of general cough in the primary model), fatigue, muscle pain, rhinorrhea, diarrhea, nausea or vomiting, chills, confusion and reporting on presence of specific prior medical conditions separately (as opposed to the presence of *any* of the prior medical conditions in the primary model).

### Baseline Models

To assess the contribution of reported symptoms and prior medical conditions to both the primary model and the extended features model (in both the Logistic Regression and the Gradient Boosting Decision Trees versions), we constructed baseline models using only age group and gender information to predict our outcome.

### Analysis of feature contributions

To gain insight into the features that contribute most to the predicted probability of being diagnosed with COVID-19 of our models, we analyzed feature contribution in the Gradient Boosting Decision Trees models using SHAP (SHapley Additive exPlanation) (10). SHAP aims to interpret the output of a machine learning model by estimating the Shapley value of each feature, which represents the average change in the output of the model, by conditioning on that feature while introducing other features one at a time, over all possible features ordering. Analyzing feature contributions in each of the models allowed us to compare the inner workings of each model and to identify which features dominated in each prediction.

We further analyzed SHAP interaction values, which uses the ‘Shapely interaction index’ to capture local interaction effects between features (10). Interaction values are calculated for each pair of the model’s features, and for each individual prediction of the model, allowing us to uncover interaction patterns between pairs of features. We placed particular emphasis on the contribution of participants’ age and the interaction of age with all other features.

## Results

Our primary model for prediction of a positive COVID-19 test result, which was constructed using Logistic Regression, achieved an area under the Receiver-Operating-Characteristic (auROC) of 0.737 (CI: 0.712-0.759), and an area under the Precision-Recall (auPR) of 0.144 (CI: 0.119-0.177). This model substantially outperforms the baseline model, which uses only age group and gender and achieved an auROC of 0.556 (CI: 0.527-0.581), and an auPR of 0.013 (CI: 0.012-0.015) (Figure 2A-B). Aside from discrimination performance measures, we also tested whether the model was calibrated. In a perfectly calibrated model, the distribution of the predicted probabilities is equal to the distribution of outcomes observed in the training data. We found that our primary model is well calibrated across the relevant prediction range (Figure 2C). The model has a positive predictive value (PPV) of 46.3%, at 10% sensitivity, while the baseline model only has 0.014%, at 10% sensitivity.

**Figure 2.**
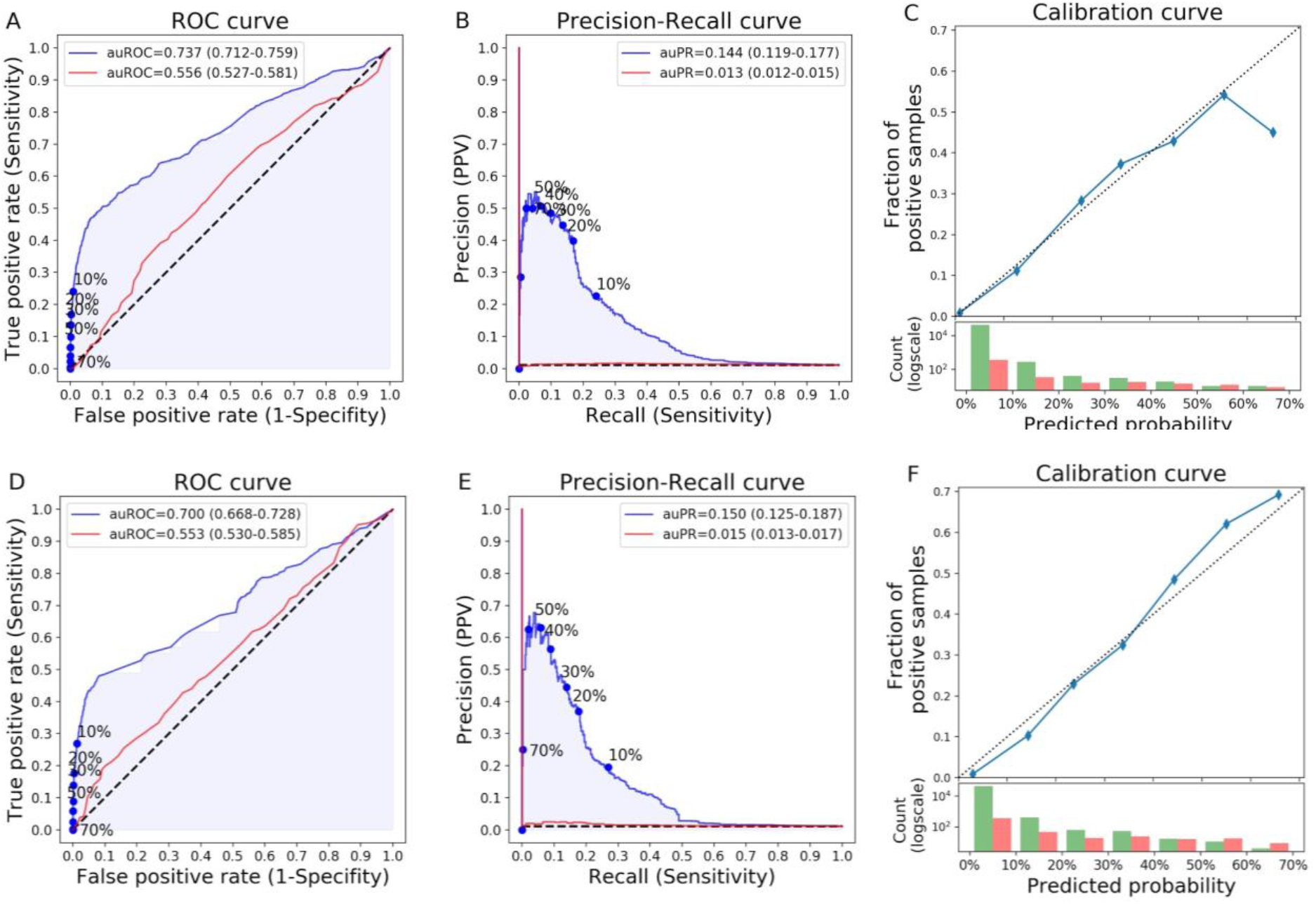
Primary model performance. **A-C:** Logistic Regression, **D-F:** Gradient Boosting Decision Trees. Abbreviations: auROC/auPR - area under the ROC/PR curve, ROC - Receiver-Operator-Characteristic, PR - Precision-Recall. Parenthesis - Confidence Interval. **A, D:** ROC curve of our model (blue) consisting of 9 simple questions and of the baseline model consisting of only age group and gender (red). Different decision probability thresholds are marked on the curve. **B, E:** Precision-Recall curve of our model (blue) and the baseline model (red). Different decision probability thresholds are marked on the curve. **C, F:** Calibration curve. Top: Blue dots represent deciles of predicted probabilities. Dotted diagonal line represents an ideal calibration. Bottom: Log-scaled histogram of predicted probabilities of COVID-19 undiagnosed (green) and diagnosed (red).

As an additional validation for the risk scores obtained, we compared the model predictions on the online survey data that was not used in the model construction process (n=121,151), with the actual number of confirmed COVID-19 patients in Israel over time. Notably, we found that the average predicted probability of individuals to test positive for COVID-19 according to our model, is highly correlated with the number of new confirmed COVID-19 cases 4 days later (pearson r=0.90, p<10^-8^) (Figure 3). Furthermore, collaborating within our Coronavirus Census Collective (CCC) (11,12), we applied the model to an independent symptom survey dataset collected every day of the week prior to conducting PCR tests in the U.S., U.K. and Sweden (n=113,139) (11,12). The model’s performance improved each day over the study period and on entries filled on the same day as PCR tests were conducted it achieved an auROC of 0.727 (CI: 0.711-0.739), an auPR of 0.217 (CI: 0.199-0.235) and a positive predictive value (PPV) of 34.7%, at 10% sensitivity, while the baseline model only has 4.55%, at 10% sensitivity (Figure 4).

**Figure 3.**
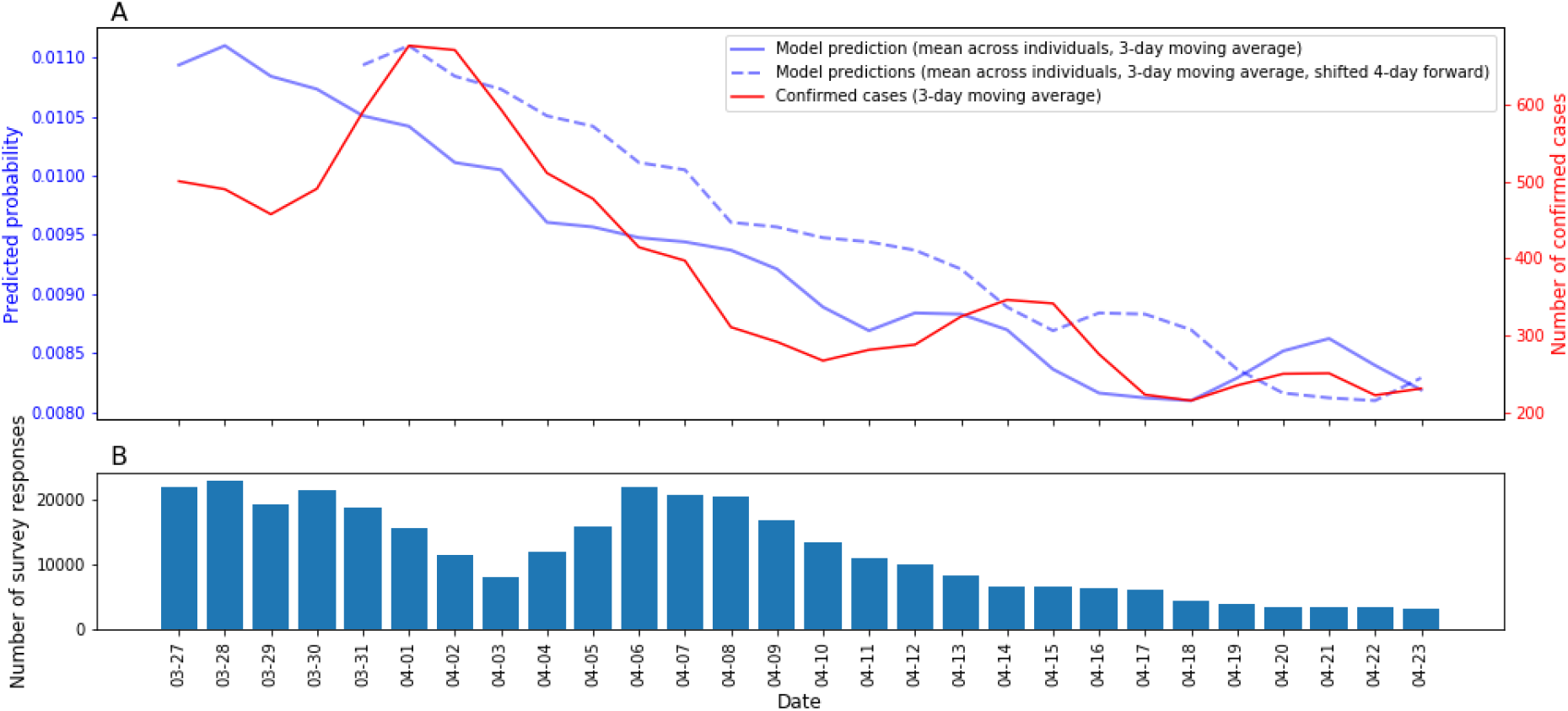
Comparison of primary model predictions to new COVID-19 cases in Israel over time. **A:** Primary model predictions, averaged across all individuals on a 3-day running average (solid blue), and shifted 4 days forward (dotted blue), compared to the number of newly confirmed COVID-19 cases in Israel by the ministry of health (MOH), based on a 3-day running average. **B:** Number of survey responses per day.

**Figure 4.**
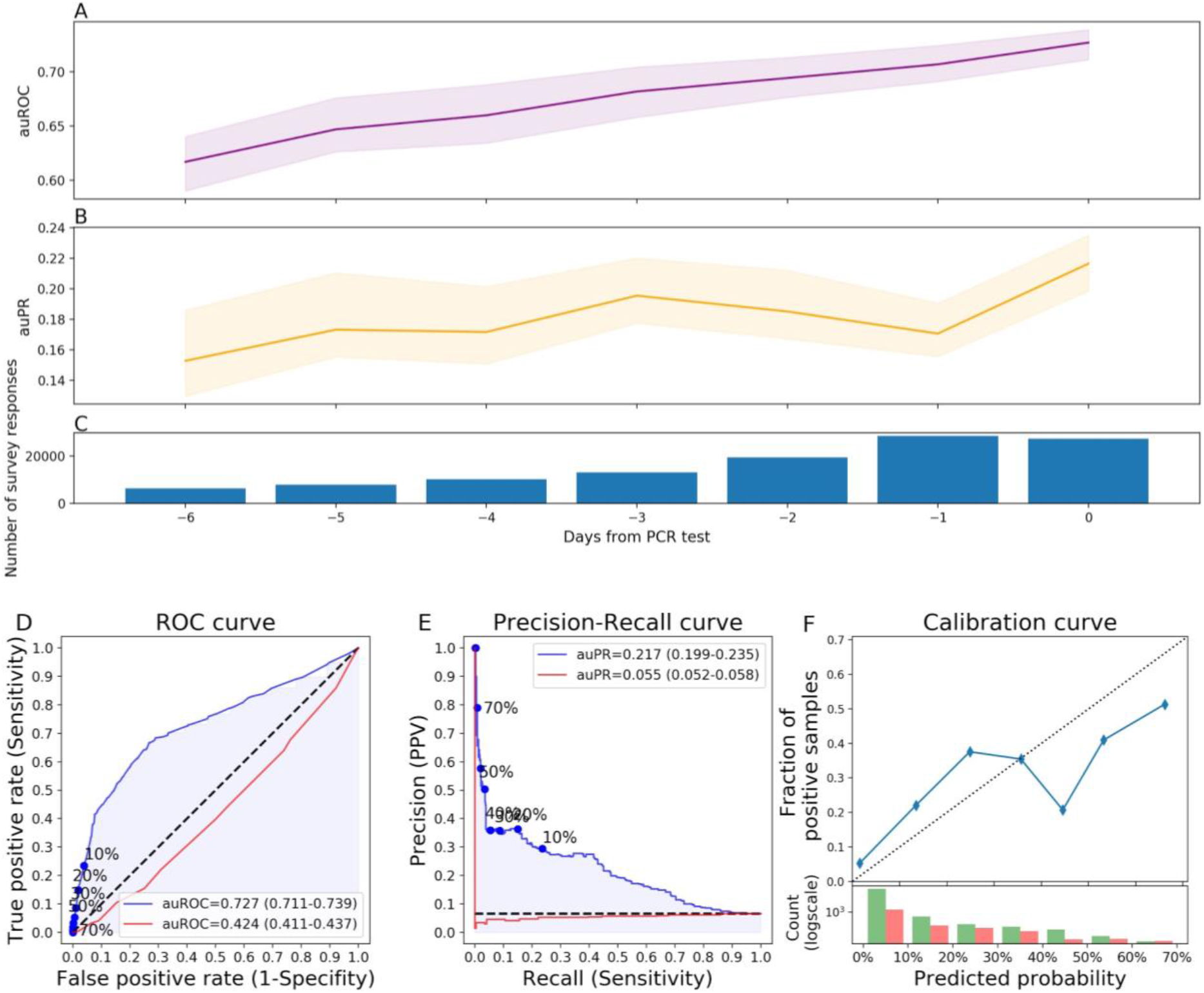
Primary model performance on independently collected dataset from the U.K, U.S. and Sweden. **A:** Area under the Receiver-Operator-Characteristic curve (auROC) (purple), **B:** Area under the Precision-Recall curve (auPR) (orange), in the days close to a PCR test. **C:** Number of survey responses per day. **D:** ROC curve of our model (blue) consisting of 9 simple questions and of the baseline model consisting of only age group and gender (red). Different decision probability thresholds are marked on the curve. **E:** Precision-Recall curve of our model (blue) and the baseline model (red). Different decision probability thresholds are marked on the curve. **F:** Calibration curve. Top: Blue dots represent deciles of predicted probabilities. Dotted diagonal line represents an ideal calibration. Bottom: Log-scaled histogram of predicted probabilities of COVID-19 undiagnosed (green) and diagnosed (red).

To better understand which features contribute to the probability of being diagnosed with COVID-19, and to examine feature interactions, we analyzed the primary model constructed using the Gradient Boosting Decision Trees algorithm (see Methods). This model showed similar performance to the Logistic Regression model (Figure 2D-F, Table S3), with a positive predictive value (PPV) of 52.6%, at 10% sensitivity, while the baseline model only has 0.025%, at 10% sensitivity. Predictions on the online survey data not used in the model’s construction process were highly correlated with the predictions of the primary Logistic Regression model (pearson r=0.91, p<10^-8^).

Analysis of feature contributions was performed using Shapley values (see Methods). Loss of taste or smell and cough had the largest overall contribution to the model (Figure 5A), when analyzing the mean absolute SHAP value of features on the entire data. Since the primary model contained a limited number of features, we compared its feature contributions to those obtained from the extended features model, also constructed using a Gradient Boosting Decision Trees algorithm. Notably, loss of taste or smell was the most contributing feature in both the primary model and the extended features model, which contained 14 additional features (Figure 5), as well as in an odds ratio analysis (Figure S1). Although the extended features model included 23 features - 14 additional features over the primary model, all symptoms included in the primary model were among the 12 features the algorithm found to be most contributing (Figure 5B).

**Figure 5.**
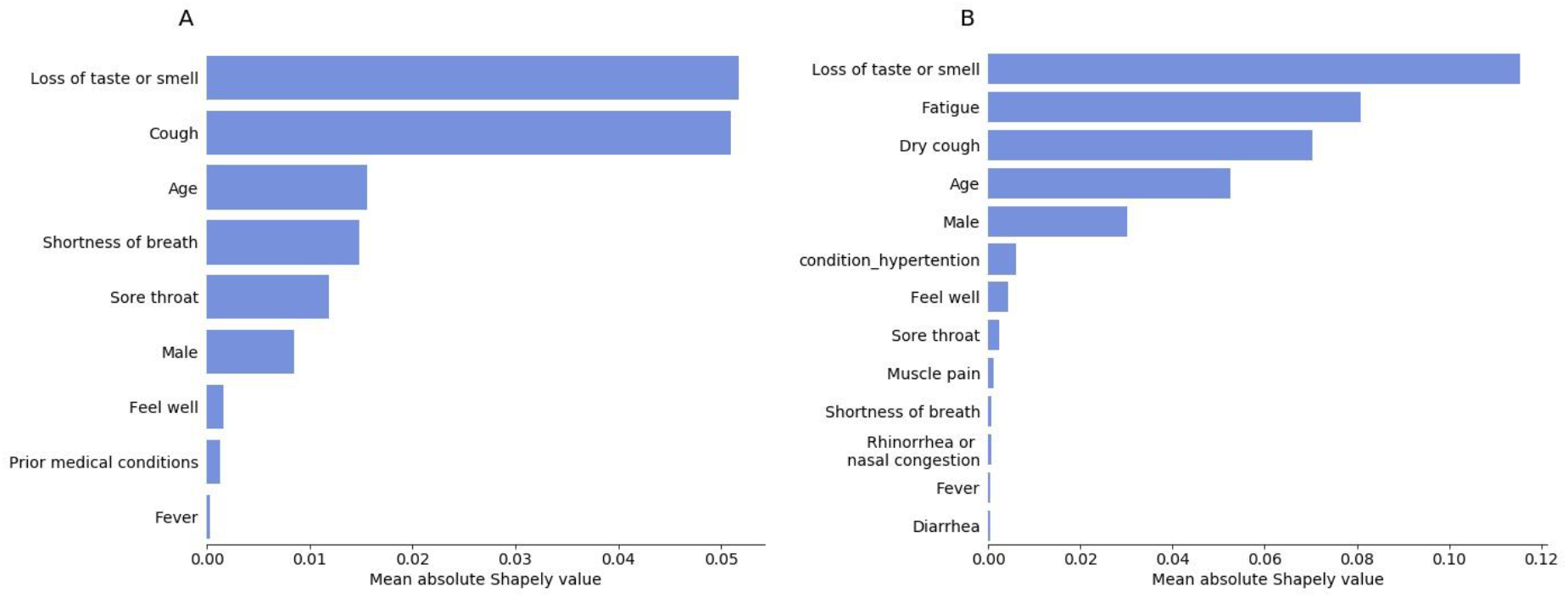
Feature contribution analysis. Mean absolute Shapley value (in units of log-odds) of **A:** the Primary model, including all features used in the model, and **B:** the Extended features model, for the 13 highest contributing features.

As age was reported as a dominant factor in COVID-19 infection and its clinical manifestation (13), we examined the interaction of age with every symptom using SHAP interaction values (see Methods). The contribution of age to the probability of being diagnosed with COVID-19 is the highest in the oldest age group (>70 years old) (Figure 6B). Presence of cough and loss of taste or smell exhibits a sharp transition-type (sigmoid-like) interaction with age, such that above the age of 40 years old, presence of each of these symptoms sharply increases the model’s predicted probability of COVID-19 infection (Figure 6G-H). In contrast, shortness of breath and sore throat show a more gradual (parabolic-like) interaction with age with presence of these symptoms increasing the model’s prediction more gradually as the age of the subject being predicted increases (Figure 6I-J). Negative answers to all these features show no interaction with age. Other examined features, such as fever and general feeling, do not show such interactions with age.

**Figure 6.**
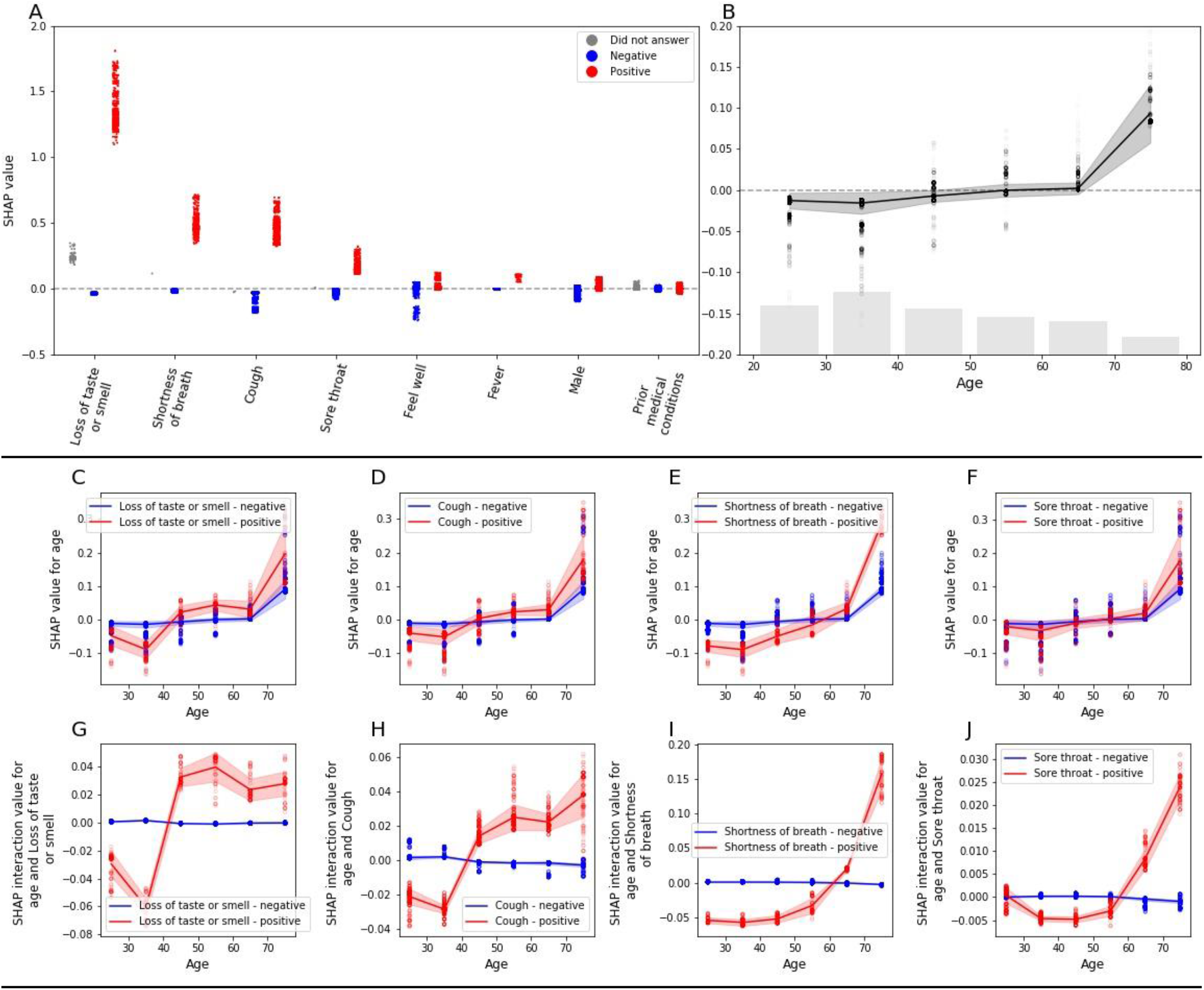
Feature interpretation analysis. **A:** SHAP values (in units of log-odds) for positive report of a feature colored in red, negative report of a feature colored in blue and missing answers in grey. **B:** SHAP values for age with number of responses as a histogram at the bottom. **C-F:** SHAP value for age, stratified by positive (red) and negative (blue) responses of loss of taste or smell (C), cough (D), shortness of breath (E) and sore throat (F). **G-J:** SHAP interaction values of age with positive (red) and negative (blue) responses of loss of taste or smell (G), cough (H), shortness of breath (I) and sore throat (J).

## Discussion

In this study we constructed a model that predicts the probability of individuals to test positive for COVID-19. Our model is based on 9 simple questions that every person can easily answer in less than a minute from the comfort of their home. Our model can assist the worldwide fight against COVID-19 by better prioritizing the limited tests available without additional costs or risk of exposure to suspected patients, thereby increasing the rate at which positive individuals can be identified and isolated.

The model was validated on a portion of the Israeli data set that was not used in the model construction process and found to be highly correlated with the number of new confirmed COVID-19 cases 4 days later. Moreover, collaborating within our Coronavirus Census Collective (CCC) (11,12), the model was validated on an independent data set collected each day in the week prior to PCR tests performance in the U.S., U.K. and Sweden. The model’s performance improved each day of the week prior to the PCR tests, probably due to the rise in symptoms and their severity in the early stages of the disease (14), while infection has yet to be confirmed and treatment has not initiated. On the day of the PCR test the model achieved a positive predictive value (PPV) of 34.7%, at 10% sensitivity, highlighting its applicability outside of Israel.

In Israel, as well as in many other countries, due to limited testing resources, suspected patients are only tested if they were exposed to a COVID-19 confirmed patient as well as exhibited acute respiratory symptoms (3). By taking an unbiased approach to predicting COVID-19 diagnosis from symptoms data, our analysis highlights the importance of additional symptoms. Of note, anosmia and ageusia that were less described in patients in the early stages of the COVID-19 pandemic (4,15) were the most impactful features in both models for predicting COVID-19 diagnosis as well as in an odds-ratio analysis. This is in line with recent literature demonstrating the importance of these symptoms in early detection and identification of the disease (16,17)^,(16,18)^. Our model also successfully recapitulated patterns of the disease that are described in the literature, such as its complex relationship with age (13). In addition, our model unraveled several patterns that are not described in the literature, such as the different patterns of interactions that particular symptoms have with age, suggesting variation of the clinical manifestation in different age groups. Although our analysis is purely predictive and not causal, these new patterns may be used to devise better testing policies, and pave the way for future studies that can uncover new aspects of the disease that were not studied to date.

Analysis of an extended features model that included 23 features compared to 9 in the primary model, validated our choice of questions in the shortened version of the survey and suggested that fatigue should also be considered. In addition, the extended features model suggested that while dry cough has an essential role in predicting COVID-19 diagnosis, moist cough does not and thus may help distinguish between cases of COVID-19 and other infections. Some of the most contributing symptoms to the prediction of a COVID-19 diagnosis are currently not included in the Israeli testing policy (3), such as loss of taste or smell and sore throat. Our analysis suggests that adding these symptoms to the testing policy may help discriminate which individuals should be tested, and improve testing prioritization.

While informative, our feature contribution analysis has several limitations. First, we did not include children in our datasets and thus, symptoms such as nausea or vomiting and diarrhea that were mostly described in children (7,8), may have a more significant part in models designed for younger age groups. Second, although we included a large set of prior medical conditions that may have a role in COVID-19 susceptibility, some of these conditions are not highly prevalent in our dataset and their contribution may thus be underestimated in our model. Finally, body temperature was the only non-mandatory question in our survey, and may thus have higher predictive power than portrayed within our model.

Several studies attempted to simulate and predict different aspects of COVID-19, such as hospital admissions, diagnosis, prognosis and mortality risk, using mostly age, body temperature, medical tests and symptoms (19). Most diagnostic models published to date were based on datasets from China and included complex features that had to be extracted through blood tests and imaging scans (19). In this work, we devised a prediction model which was based solely on self-reported information, and as such it could be easily deployed and used instantly in other countries.

Our study has several additional limitations. First, our data is biased by Israel’s MOH ever changing testing policy, such that at some point all of the COVID-19 positively diagnosed participants in our study had to be eligible for a test under that policy. An ideal dataset for purposes of devising a classifier should include a large random sampling of the population, but such data coupled with symptom surveys is currently unavailable at large-scale. Accordingly, all the diagnosed responders in our study are not in the first stage of showing symptoms, but are in some time-lag after diagnosis. In addition, our study is based on self reports of willing participants and is therefore bound to suffer from some selection bias. The bias is significantly reduced in the data collected via the IVR platform, since all residents in the IVR-surveyed cities were actively contacted only once, on the same day and in the same manner. In the online version of the survey we made attempts to reduce this bias by promoting it in several media outlets and by engaging leaders of underrepresented communities. Finally, we acknowledge that the ideal choice of a baseline model would be the Israeli MOH testing policy, but since it constantly changed during the pandemic and included questions that were not part of our survey we were unable to compare to it. Nevertheless, at an unknown sensitivity (as we do not know the actual number of cases), the overall predictive value (PPV) of all tests in Israel was 4.6% (3), and although the measures are not directly comparable, we believe that our model substantially outperforms this PPV.

In conclusion, our constructed model predicts COVID-19 PCR test results with high discrimination (positive predictive value (PPV) of 46.3% at a 10% sensitivity) and calibration. It also suggests that several symptoms that are currently not included in the Israeli testing policy exhibit intriguing interactions with age and should probably be integrated into revised testing policies. Overall, our approach can be utilized worldwide to direct the limited resources towards individuals who are more likely to test positive for COVID-19, leading to faster isolation of infected patients and therefore to reduced rates of virus spread.

## Data Availability

Models and tables of de-identified, aggregated data are available at https://github.com/hrossman/Covid19-Survey.

## Code availability statement

Source code for the survey is available at https://github.com/hasadna/avid-covider as an open source project, and can be readily adapted to use in other countries.

## Ethics Declarations

The study protocol was approved by the Weizmann Institute of Science review board (IRB). Informed consent was waived by the IRB, as all identifying details of the participants were removed before the computational analysis. Participants were made fully aware of the way in which the data will be stored, handled and shared, which was provided to them and is in accord with the privacy and data-protection policy of the Weizmann Institute of Science (https://weizmann.ac.il/pages/privacy-policy).

## Competing Interests Statement

The authors declare no competing interests.

## Authors contribution

S.S, T.K., A.K., S.S. and H.R. conceived the project, designed and conducted the analyses, interpreted the results and wrote the manuscript. A.G., T.M., A.L., D.K., I.K., A.G. designed and conducted the survey. O.C. directed the organizational and logistic efforts. A.K., O.H., M.Z.A., N.C., A.E.Z., A.I. designed and conducted the survey as well as provided the data. B.G., D.H., V.S., R.B. and E.S. conceived, directed and supervised the project and analyses.

## Acknowledgments

We thank the following for their contributions to our efforts: Y. Landau, T. Eldar, S. Kasem, T. Bria, S. Avraham, B. Kirel, A. Terkeltaub, N. Kalkstein, E. Krupka, M. Loitsker, Y. Sela, T. Bareia, J. Shalev, A. Brik, L. Samama, S. Cohen, J. Bitton, R. Priborkin, G. Beryozkin, G. Dardyk, S. Lanzman, N. Epstein, N. B. Shizaf, E. Daian, M. Bialek, M. Suleiman, R. Maor, Z. Maor, Y. Dinur, J. Klinger, R. Zaida, M. Eden, N. Hirshman, O. Dovev, Y. Grossman and the Public Knowledge Workshop (‘Hasadna’).

---

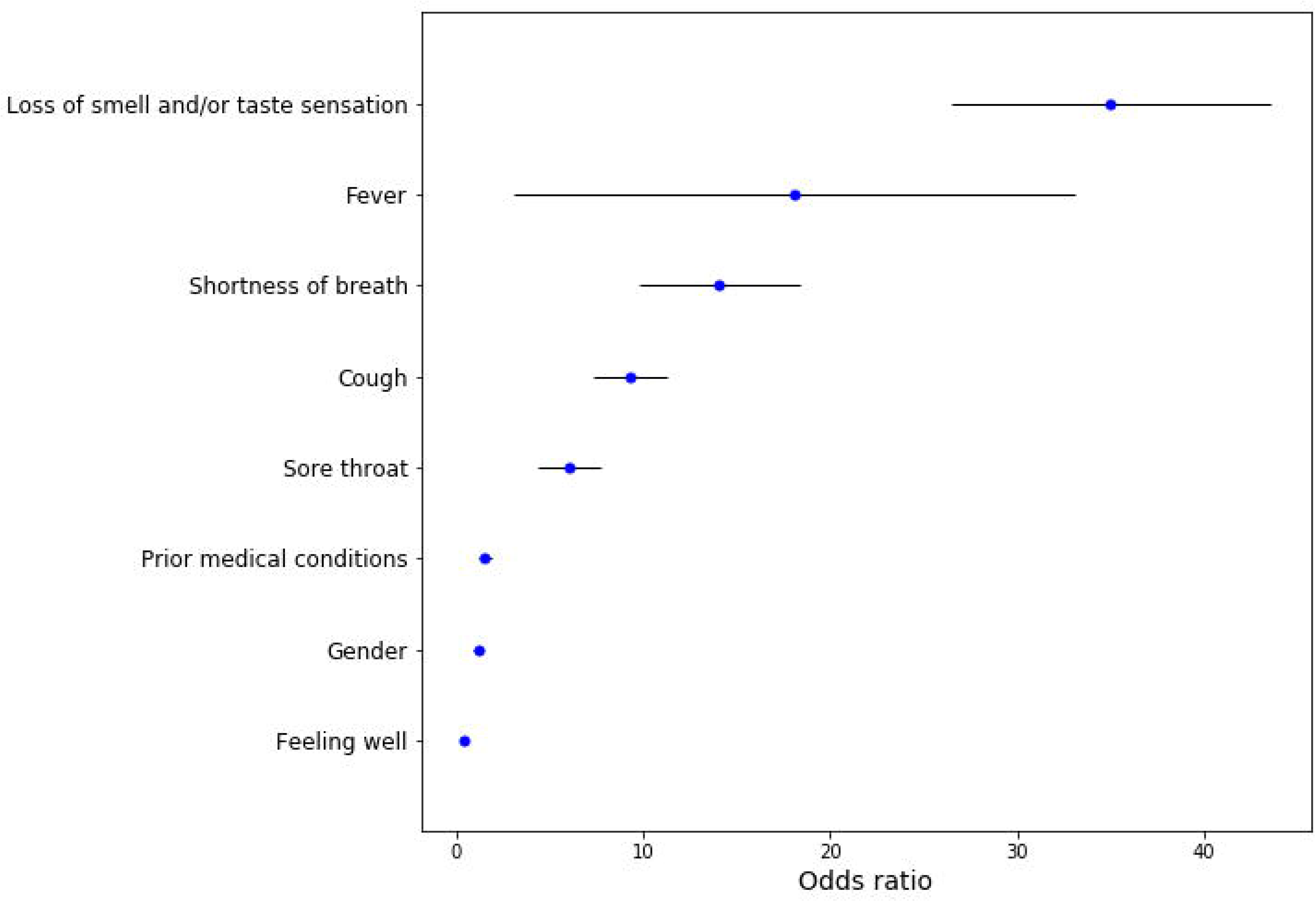

## Notes

### Competing Interest Statement

The authors have declared no competing interest.

### Funding Statement

no external funding was received

